# Comparison of MIS-C Related Myocarditis, Classic Viral Myocarditis, and COVID-19 Vaccine related Myocarditis in Children

**DOI:** 10.1101/2021.10.05.21264581

**Authors:** Trisha Patel, Michael Kelleman, Zachary West, Andrew Peter, Matthew Dove, Arene Butto, Matthew E. Oster

## Abstract

**Background:** Although rare, myocarditis in the pediatric population is a disease process that carries significant morbidity and mortality. Prior to the SARS-CoV-2 related (COVID-19) pandemic, enteroviruses were the most common cause of classic myocarditis. However, since 2020, myocarditis linked to multisystem inflammatory syndrome in children (MIS-C) is now common. In recent months, myocarditis related to COVID-19 vaccines has also been described. This study aims to compare these three different types of myocarditis with regards to clinical presentation, course, and outcomes.

**Methods:** In this retrospective cohort study, we included all patients <21 years of age hospitalized at our institution with classic viral myocarditis from 2015-2019, MIS-C myocarditis from 3/2020-2/2021 and COVID-19 vaccine-related myocarditis from 5/2021-6/2021. We compared demographics, initial symptomatology, treatment, laboratory data, and echocardiogram findings.

**Results:** Of 201 total participants, 43 patients had classic myocarditis, 149 had MIS-C myocarditis, and 9 had COVID-19 vaccine-related myocarditis. Peak troponin was highest in the classic myocarditis group, whereas the MIS-C myocarditis group had the highest recorded brain natriuretic peptide (BNP). There were significant differences in time to recovery of normal left ventricular ejection fraction (LVEF) for the three groups: nearly all patients with MIS-C myocarditis (n=139, 93%) and all patients with COVID-19 vaccine-related myocarditis (n=9, 100%) had normal LVEF at the time of discharge, but a lower proportion of the classic myocarditis group (n=30, 70%) had a normal LVEF at discharge (p<0.001). Three months post-discharge, 18 of 40 children (45%) in the classic myocarditis group still required heart failure treatment, whereas only one of the MIS-C myocarditis patients and none of the COVID-19 vaccine-associated myocarditis patients did.

**Conclusions:** Compared to those with classic myocarditis, those with MIS-C myocarditis had more significant hematologic derangements and worse inflammation at presentation, but had better clinical outcomes, including rapid recovery of cardiac function. Patients with COVID-19 vaccine-related myocarditis had similar clinical presentation to patients with classic myocarditis, but their pattern of recovery was similar to those with MIS-C, with prompt resolution of symptoms and improvement of cardiac function. Long-term follow-up should focus on cardiac and non-cardiac consequences of myocarditis associated with COVID-19 illness and vaccination.

## Introduction

Myocarditis is an inflammatory disease of the myocardium with a variety of causes and a spectrum of presentations, ranging from subclinical disease to acute fulminant myocarditis that leads to cardiogenic shock.^1^ In the pediatric population, myocarditis is rare with an approximate incidence of 0.05% and is most commonly virally-mediated, with high prevalence of detection of enterovirus, adenovirus, parvovirus B19, and Ebstein-Barr virus (EBV).^1, 2^ Regardless of initial presentation, some patients with so-called “classic” myocarditis have full recovery of cardiac function, while others may develop residual cardiomyopathy and chronic congestive heart failure. A recent single center study demonstrated that within a population of 41 pediatric patients with myocarditis, 66% made full recovery, 10% had incomplete recovery of function, and 24% either died or underwent transplantation.^3^ Long-term follow-up in the pediatric population of classic myocarditis is limited, but myocarditis has been shown to be the cause of dilated cardiomyopathy in 27-46% of newly diagnosed cases.^4, 5^

Although classic viral myocarditis was the most common cause of myocarditis in children prior to 2020, since the onset of the SARS-CoV-2 related (COVID-19) pandemic, multisystem inflammatory syndrome in children (MIS-C) linked to severe acute respiratory syndrome coronavirus 2 (SARS-CoV-2) has proven to be another important cause of myocarditis. In children with MIS-C, myocarditis has been found to be a common complication, with an estimated incidence of 17-75% in affected children.^6-8^ Little is currently known about the long term cardiac sequalae of MIS-C myocarditis, specifically in regards to improvement in cardiac function, given the novelty of the disease. Existing studies evaluating changes to cardiac function in this patient population over time are limited but have shown near-complete recovery of left ventricular ejection fraction (LVEF) in most children with MIS-C myocarditis. ^9-11^

In recent months, a new etiology of myocarditis related to the mRNA-based COVID-19 vaccines has been proposed in the pediatric population. This type of myocardial injury has typically presented in adolescent and young adult males, most commonly after a second dose of the vaccine. ^12-14^ Information about this new entity is rapidly evolving and little is known about how this vaccine-mediated myocarditis differs from classic myocarditis.

These three types of myocarditis appear to be distinct and little is currently known about the differences between these etiologies. We aimed to compare findings and outcomes amongst classic myocarditis, MIS-C myocarditis, and COVID-19 vaccine-related myocarditis.

## Methods

In this retrospective cohort study, we evaluated patients <21 years of age with classic myocarditis, MIS-C myocarditis, and COVID-19 vaccine-related myocarditis at our institution. The classic myocarditis group consisted of patients who were diagnosed with viral or idiopathic myocarditis from 2015-2019. The MIS-C myocarditis cohort included patients from March 2020-February 2021 who met CDC case definition of MIS-C and had evidence of myocardial involvement with troponin elevation. ^15^ The COVID-19 vaccine-related group consisted of patients from May 2021-June 2021 with myocarditis after the vaccine, with no other etiology of myocarditis identified. The study was approved by the Institutional Review Board of Children’s Healthcare of Atlanta.

For eligible patients, key outcomes of interest included LVEF at the time of presentation, discharge, and at clinic follow-up; peak troponin I and brain natriuretic peptide (BNP) levels; and medical treatment for myocarditis. Other variables of interest were symptomatology at presentation, electrocardiographic (ECG) findings, and laboratory evidence of anemia, thrombocytopenia, and lymphopenia. Additionally, clinical status at two time points, time of hospital discharge and three-month follow-up after initial presentation, were included when available. Clinical status outcomes included follow-up without oral heart failure medications, follow-up with oral heart failure medications, left ventricular assist device (LVAD) placement, and transplant or death. Oral heart failure medications included diuretics, angiotensin-converting enzyme (ACE) inhibitors, angiotensin II receptor blockers (ARB), aldosterone antagonists, and beta blockers.

### Statistical Analysis

Statistical analyses were performed using SAS v9.4 (SAS Institute, Cary, NC) and statistical significance was assessed at the 0.05 level. Normality of continuous variables was assessed using histogram, normal probability plots, and the Anderson-Darling test for normality. Descriptive statistics are presented as counts and percentages for categorical variables and median (25th percentile – 75th percentile) for continuous data with skewed distributions.

Continuous data were compared between myocarditis groups using Kruskall-Wallis tests and comparisons between categorical variables were performed using Chi-square tests, or Fisher’s exact test when expected cell counts were <5. The time-dependent outcome of normal LVEF was analyzed with survival analysis. Because in-hospital mortality was considered as a competing event for normal LVEF, a competing-risk analysis was performed.

After performing the overall analyses, similar measures were undertaken to do sub-analyses comparing patients ≥12 years in all three groups. These comparisons were completed given that vaccines were not authorized during the study period in those <12 years.

## Results

### Demographics and presenting symptoms

A total of 201 patients with myocarditis were included in this study, 43 of whom were in the classic myocarditis arm, 149 in the MIS-C myocarditis arm, and 9 in the vaccine-related arm (Table 1). In the vaccine-related group, eight of the nine patients presented with myocarditis after the second vaccine dose; among those patients, there was a mean of 5.4 days and median of 3 days between vaccination dose number two and hospital admission.

**Table 1:**
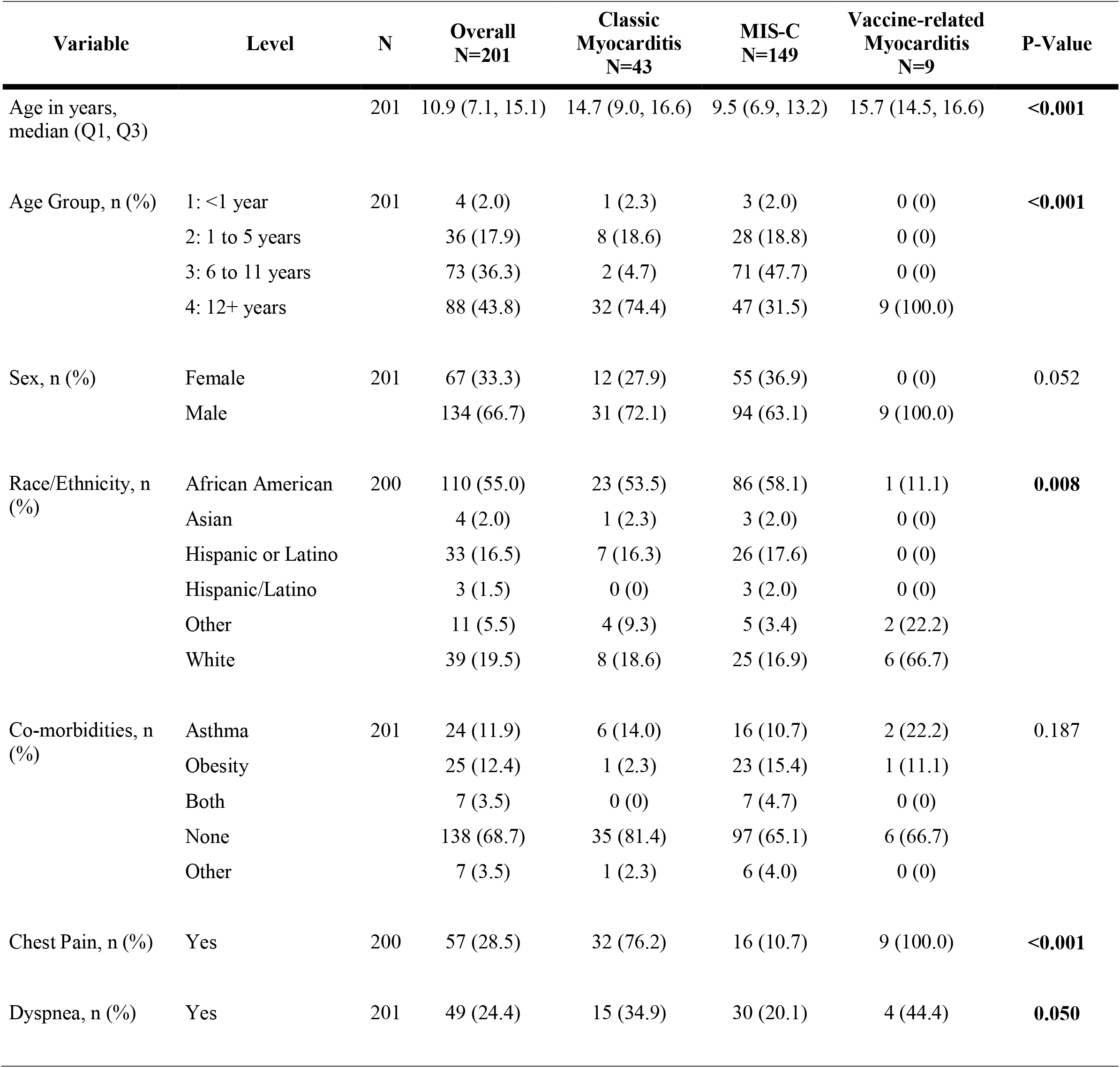
Demographics and Cardiac Symptomology and Signs Among Patients with Classic Myocarditis, MIS-C myocarditis, and Vaccine-related Myocarditis

Those with MIS-C were younger than those with classic myocarditis (median 7.5 years vs. 14.7 years) and vaccine-related myocarditis (median 15.5 years). All three groups had predominantly male sex. More than half of the children with classic myocarditis or MIS-C were of African American non-Hispanic race/ethnicity, while two thirds of the vaccine-related myocarditis group were White non-Hispanic. Among presenting symptoms, more patients with classic myocarditis and vaccine-related myocarditis presented with chest pain (76.2% and 100%, respectively) compared to patients with MIS-C myocarditis (10.7%, p<0.001).

### Laboratory findings

There were significant differences between the three groups when comparing cardiac biomarkers and inflammatory markers (Figure 1). Peak troponin was highest in the classic myocarditis group (median 16 ng/mL, interquartile range (IQR) 8.1 to 30.6), while the MIS-C myocarditis group had the highest recorded BNP (median 932 pg/mL, IQR 459 to 1672). The MIS-C group also had the most prominent hematologic derangements, including lymphopenia (median 0.6 × 10^3^/uL, IQR 0.4 to 0.99) with leukocytosis (median 15.1 × 10^3^/uL, IQR 11 to 20), thrombocytopenia (median 145 × 10^3^/uL, IQR 110 to 189), and anemia (median 9.4 g/dL, IQR 8.6 to 10.4).

**Figure 1:**
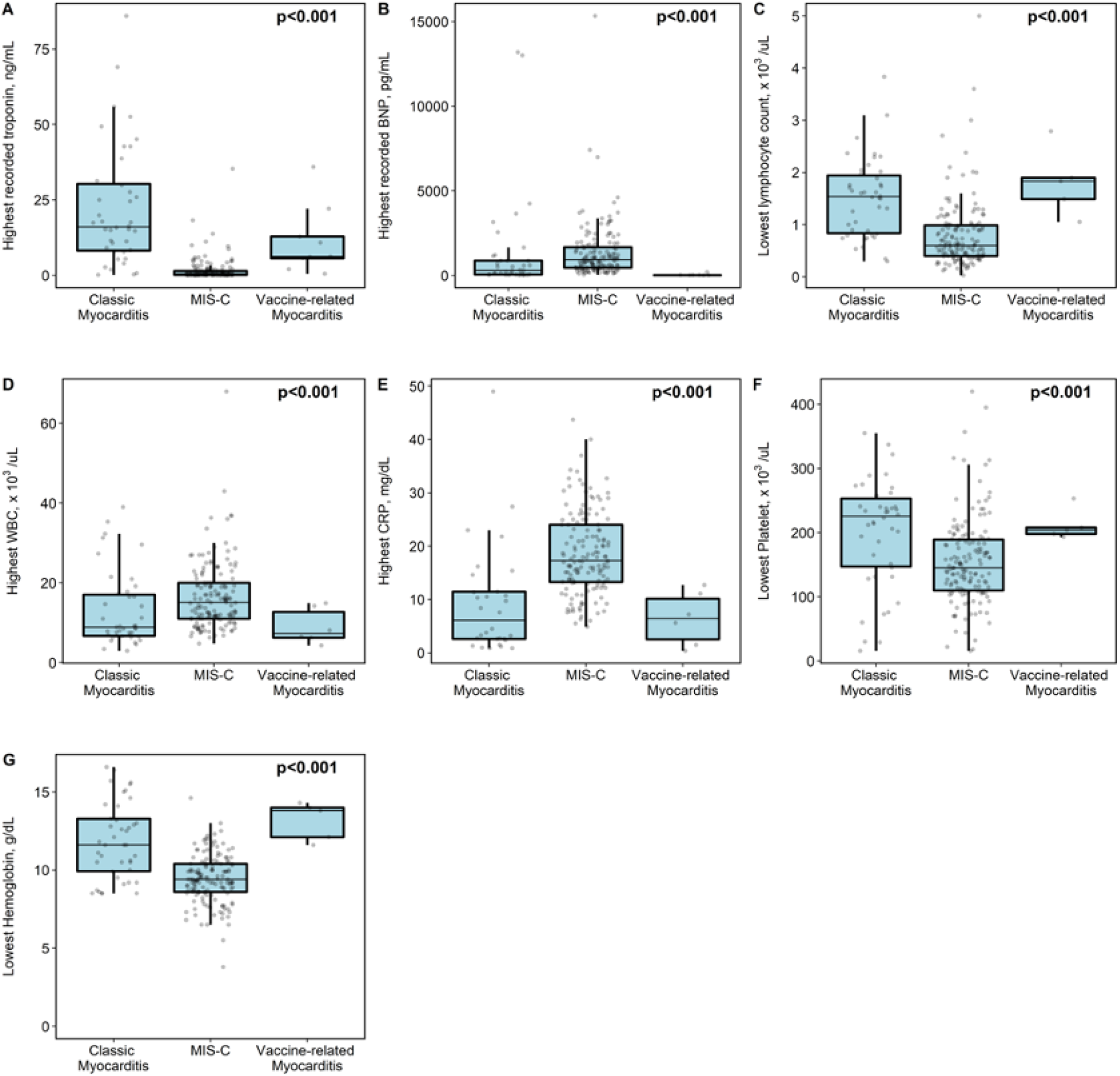
Comparison of laboratory testing among those with classic myocarditis, multisystem inflammatory syndrome in children (MIS-C), or COVID-19 vaccine-related myocarditis. Peak troponin was highest in the classic myocarditis group, whereas the MIS-C myocarditis group had highest recorded brain natriuretic peptide (BNP), most profound lymphopenia, highest white blood cell count (WBC), highest C-reactive protein (CRP), lowest platelet level, and lowest hemoglobin level.

### Electrocardiogram and Echocardiogram Findings

ECG abnormalities were common in those with classic myocarditis (74%) and vaccine-related myocarditis (67%), but fewer than half of children with MIS-C myocarditis had ECG abnormalities (Table 2). The most common ECG abnormality was a repolarization abnormality, present in 58% of those with classic myocarditis, 18.5% of those with MIS-C, and 67% of those with vaccine-related myocarditis.

**Table 2:** Cardiac Testing Among Patients with Classic Myocarditis, MIS-C myocarditis, and Vaccine-related Myocarditis Myocarditis

| Variable | Level | N | Overall<br>N=201 | Classic<br>Myocarditis<br>N=43 | MIS-C<br>N=149 | Vaccine-related<br>Myocarditis<br>N=9 | P-Value |
| --- | --- | --- | --- | --- | --- | --- | --- |
| Left ventricular ejection fraction at presentation, categorized, n (%) | 01: <30% | 201 | 7 (3.5) | 4 (9.3) | 3 (2.0) | 0 (0) | 0.066 |
|  | 02: 30-55% |  | 82 (40.8) | 21 (48.8) | 59 (39.6) | 2 (22.2) |  |
|  | 03: >55% |  | 112 (55.7) | 18 (41.9) | 87 (58.4) | 7 (77.8) |  |
| Left ventricular ejection fraction at presentation, median (Q1, Q3) |  | 201 | 56 (46, 64) | 51.0 (38.0, 61.0) | 56.2 (48, 64.8) | 59.7 (57.8, 67.2) | **0.019** |
| Lowest recorded EF, median (Q1, Q3) |  | 201 | 50.2 (39, 58.6) | 45.3 (29.4, 60) | 50.2 (40.0, 57.6) | 59.7 (57.8, 67.2) | **0.013** |
| Normal EF at the time of discharge, n (%) | Yes<sup>1</sup> | 201 | 178 (88.6) | 30 (73.2) | 139 (93.3) | 9 (100.0) | **<0.001** |
| Coronary artery dilation at time of presentation, n (%) | No | 199 | 186 (93.5) | 42 (97.7) | 135 (91.8) | 9 (100.0) | 0.440 |
| Coronary artery dilation at discharge, n (%) | No | 199 | 191 (96.0) | 41 (97.6) | 141 (95.3) | 9 (100.0) | 0.786 |
| Worst pericardial effusion type, n (%) | Absent | 201 | 102 (50.7) | 22 (51.2) | 72 (48.3) | 8 (88.9) | **0.005** |
|  | Present |  | 97 (48.3) | 21 (48.8) | 76 (51.0) | 0 (0) |  |
|  | Trivial |  | 2 (1.0) | 0 (0) | 1 (0.7) | 1 (11.1) |  |
| ECG findings at admission, n (%) | Normal | 198 | 125 (63.1) | 12 (27.9) | 110 (73.8) | 3 (33.3) | **<0.001** |
|  | Complete AV block |  | 3 (1.5) | 3 (7.0) | 0 (0) | 0 (0) |  |
|  | Repolarization abnormalities |  | 58 (29.3) | 25 (58.1) | 27 (18.5) | 6 (66.7) |  |
|  | Other<sup>2</sup> |  | 12 (6.1) | 3 (7.0) | 9 (6.2) | 0 (0) |  |
| ECG findings at<br>clinic follow up (1-2<br>weeks after<br>discharge), n (%) | Normal | 183 | 168 (91.8) | 29 (74.4) | 134 (97.1) | 5 (83.3) | <0.001 |
|  | Repolarization<br>abnormalities |  | 5 (2.7) | 3 (7.7) | 1 (0.7) | 1 (16.7) |  |
|  | Other<sup>2</sup> |  | 10 (5.5) | 7 (17.9) | 3 (2.2) | 0 (0) |  |

Abbreviations: EF (ejection fraction), ECG (electrocardiogram), AV (atrioventricular)
<sup>1</sup> The EF at the time of discharge for two additional patients with classic myocarditis, one who died and one who progressed to transplant, are not included in this row of the table
<sup>2</sup> Other ECG findings included Brugada pattern, atrial tachyarrhythmia, markedly abnormal ventricular axis, junctional tachycardia, and abnormal intervals (i.e. prolonged QTc and PR)

On echocardiogram, there were notable differences in LVEF and in the presence of pericardial effusion between groups, but there was no significant difference in the incidence of coronary artery dilation (Table 2). At presentation, LVEF was lowest for those with classic myocarditis, with EF <55% present in 58% of patients, compared to 42% of MIS-C myocarditis and 22% of vaccine-related myocarditis. Patients with classic myocarditis also had the lowest recorded EF (median 45%, IQR 29-60%). Among those with transplant-free survival at the time of discharge, 26% of classic myocarditis patients and 7% of MIS-C patients had EF <55%; all patients with vaccine-related myocarditis patients had EF >55% at discharge. Pericardial effusion was present in roughly half of the classic and MIS-C myocarditis patients (49% and 52%, respectively), but was rare in the vaccine-related myocarditis patients (one patient with trivial pericardial effusion).

### Treatment

Treatment of myocarditis varied depending on the type (Table 3). In patients with MIS-C myocarditis, intravenous immunoglobulin (IVIG) and steroids were frequently used, in accordance with our hospital protocol for MIS-C management.^16^ IVIG was commonly used in classic myocarditis (65%), but in only one patient with vaccine-related myocarditis. Steroids were occasionally used in classic myocarditis (33%), and were not used at all in vaccine-related myocarditis. Vasopressors were most often used in those with classic myocarditis (42%) or MIS-C (52%), and rarely used in vaccine-related myocarditis (22%). Patients with classic myocarditis were more likely to require mechanical ventilation (28%), compared with only 4% of MIS-C patients. ECMO (extracorporeal membrane oxygenation) was used in 21% of classic myocarditis patients and 2% (n=3) of MIS-C patients. Only two children with vaccine-related myocarditis had ICU admission; neither received mechanical ventilation or ECMO.

**Table 3:**
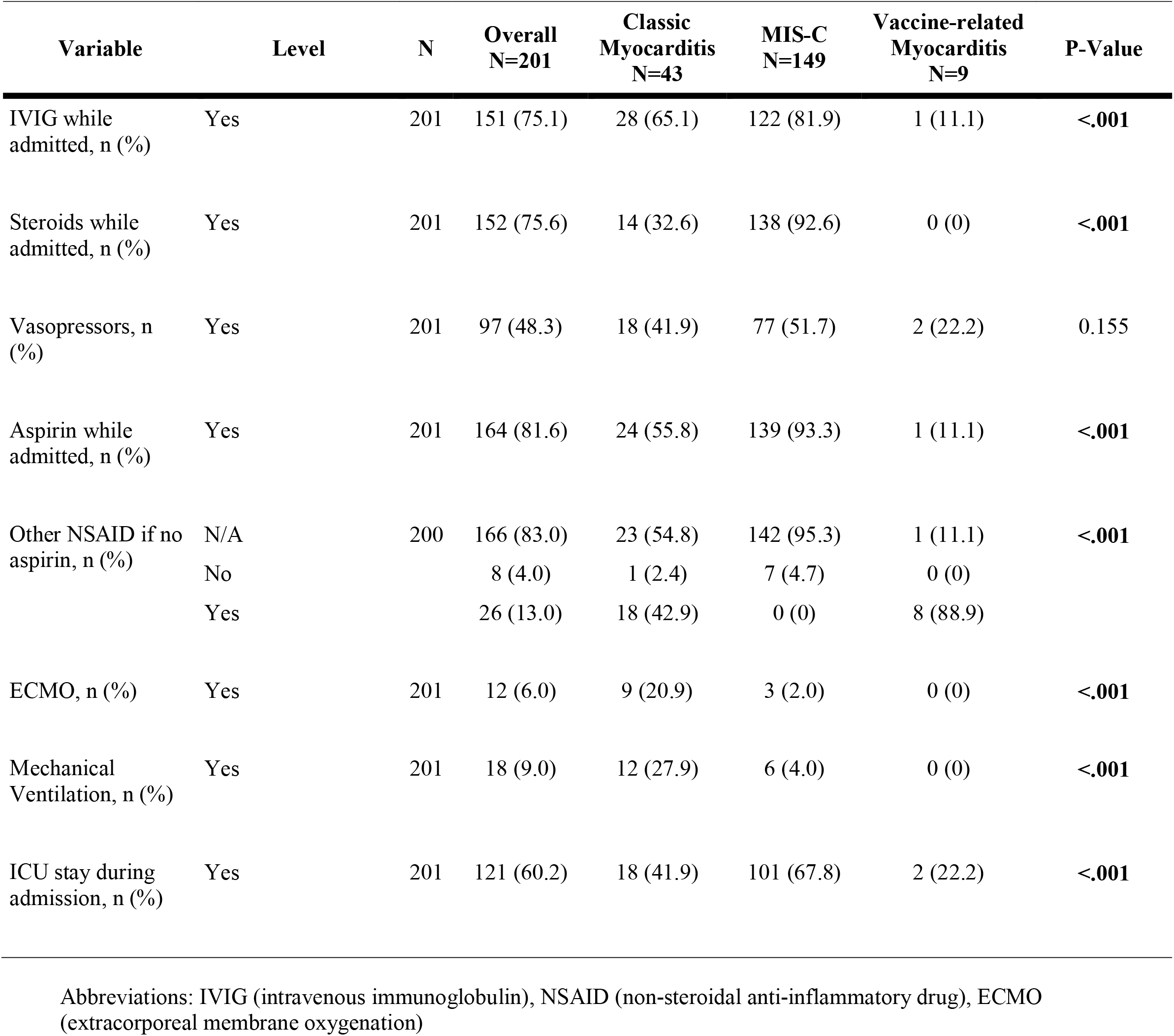
Treatment Variation Among Patients with Classic Myocarditis, MIS-C myocarditis, and Vaccine-related Myocarditis

### Outcomes

There was one death and one heart transplant among the classic myocarditis group, with all other patients in this study discharged to home (Figure 2). Approximately 44% of the classic myocarditis patients were discharged on oral heart failure medications, while only 3% of MIS-C myocarditis patients were discharged on oral heart failure therapy; no patients with vaccine-related myocarditis patients required oral heart failure therapy at discharge. These proportions were similar at 3-month follow-up (Figure 2). Of note, among the 21 patients with classic myocarditis and MIS-C myocarditis that were discharged from the hospital with depressed function, all 10 of the MIS-C myocarditis patients had ventricular recovery three months after discharge, while 3 of the 11 (27%) patients with classic myocarditis had persistent ventricular dysfunction.

**Figure 2:**
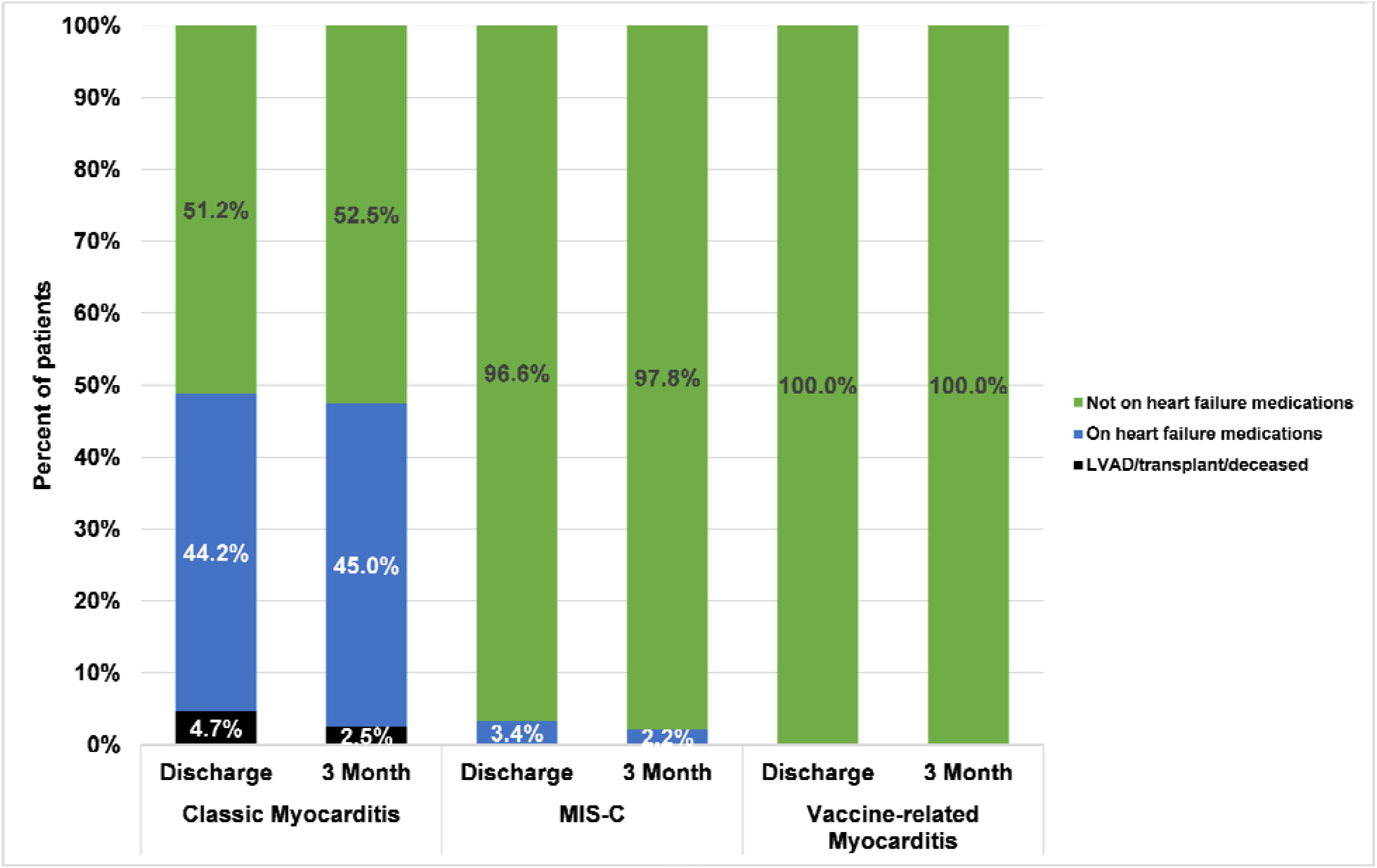
Comparison of discharge outcomes and three month outcomes among those with classic myocarditis, multisystem inflammatory syndrome in children (MIS-C), or COVID-19 vaccine-related myocarditis.

Timing of ventricular recovery varied among the three groups (Figure 3). Within three days of hospitalization, only 47% of those with classic myocarditis had regained normal function (EF >55%), while 76% of those with MIS-C myocarditis and all of those with vaccine-related myocarditis had achieved that target. Within 10 days of presentation, 70% of those with classic myocarditis and 94% of those with MIS-C had regained normal cardiac function.

**Figure 3:**
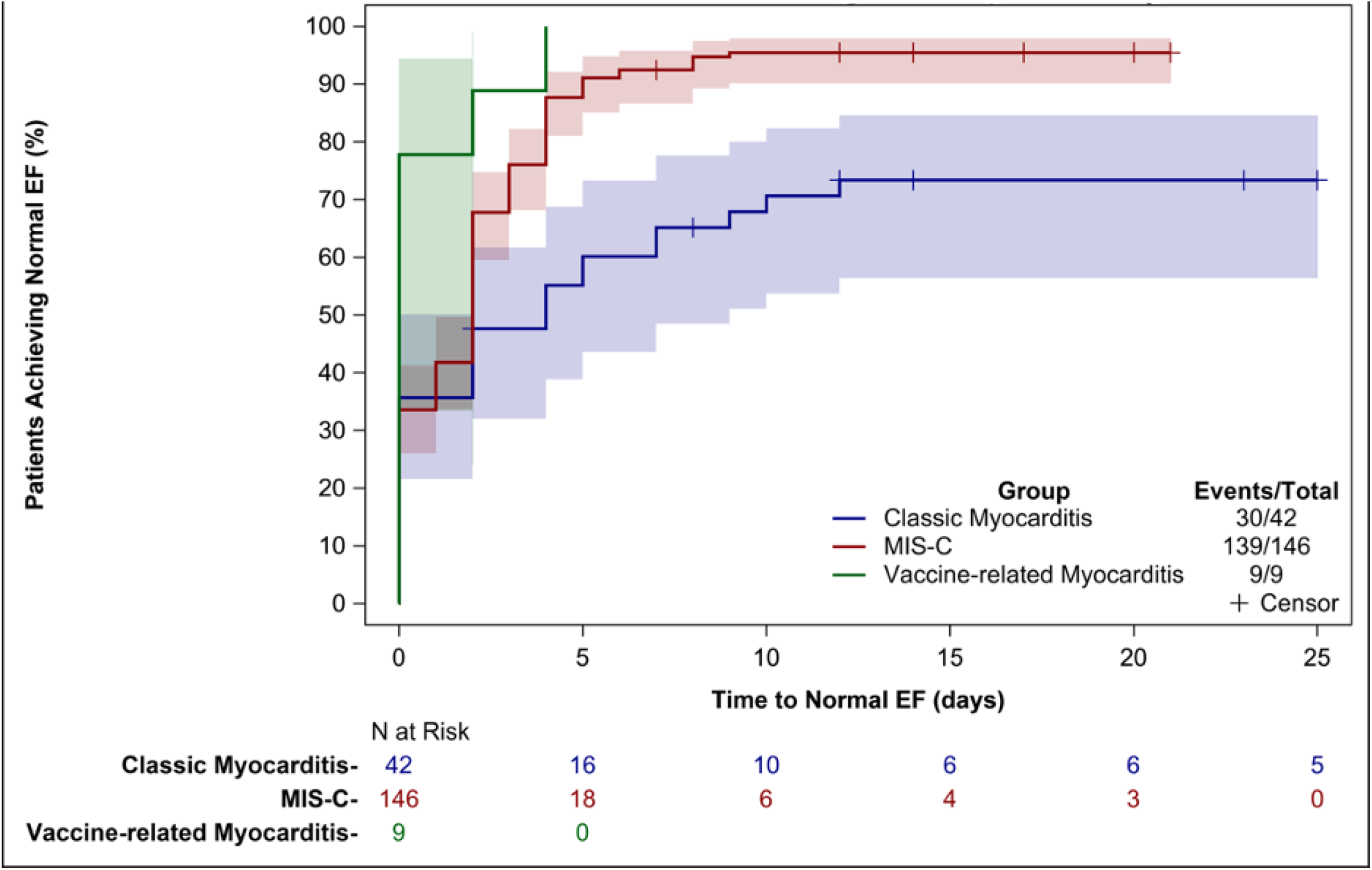
Cumulative incidence function accounting for in-hospital mortality

### Subanalyses (Supplemental tables)

When restricting the analysis to children who were ≥ 12 years, we found similar findings to the total cohort. Notable differences included higher rates of co-morbidities and greater need for vasoactive support in the older children with MIS-C when compared to the two other groups (Supplemental Table 1, Supplemental Table 2, Supplemental Table 3, Supplemental Figure 1). Differently than the full cohort, in patients ≥ 12 years, the MIS-C patients tended to have the lowest recorded median EF. The majority of affected MIS-C patients had full recovery of cardiac function at the time of discharge, similar to the analysis of children of all ages (Supplemental Table 2).

## Discussion

To our knowledge, this is the first analysis comparing findings in patients with classic myocarditis, MIS-C myocarditis, and COVID-19 vaccine-related myocarditis. Compared to classic myocarditis, patients with MIS-C had a greater likelihood of full recovery of cardiac function with a faster time to recovery, even when they presented with fulminant myocarditis. Those with COVID-19 vaccine-related myocarditis generally had a milder clinical course, with lower likelihood of cardiac dysfunction at presentation and more rapid recovery when present.

Similar to our study, other analyses have also highlighted the return of normal systolic cardiac function in the follow up of patients with MIS-C myocarditis.^**11**, **17**^ This ventricular recovery is a key finding and is distinct from the pattern of recovery seen in patients with classic myocarditis, in whom ventricular recovery tends to be slow. Other children in the classic myocarditis population never recovered function, progressing to heart transplantation or death. Thus, although patients with MIS-C myocarditis can present with severely depressed cardiac function requiring intensive care, mechanical ventilation, and even ECMO, reassuringly patients in the short term regain normal cardiac function and continue to have evidence of full cardiac recovery at the time of their three-month follow-up visits.

One explanation for this rapid recovery for children with MIS-C myocarditis compared to classic myocarditis is that cardiac dysfunction is a result of the systemic inflammatory state associated with MIS-C; once that has been addressed, cardiac function recovers. Recovery may be slower in classic myocarditis because of direct myocyte injury as well as immune mediation due to a viral trigger, in the case of viral myocarditis.^18^ Another factor to consider in the MIS-C population is the impact that treatment with IVIG and steroids may have had in the return to full recovery of cardiac function. There was a statistically significant difference between the MIS-C myocarditis and classic myocarditis group, in that the MIS-C group received treatment more readily when compared to the classic myocarditis group, and thus this could play a role in the difference in return to recovery. However, in the treatment of classic myocarditis, IVIG is not routinely recommended given its variable efficacy in studies so its role in the improvement of EF in the setting of MIS-C myocarditis is also unclear.^5^

In our study, MIS-C myocarditis tended to affect a younger population, similar to findings in a large-scale analysis of patients with MIS-C with a median age of 8 years.^19^

Interestingly, patients with classic myocarditis in our study tended to be older and to present with chest pain when compared to the MIS-C myocarditis group. This difference could be explained by the fact that the older patient population within the classic myocarditis group was more readily able to express chest discomfort compared to the younger population in the MIS-C myocarditis group. The COVID-19 vaccine-related myocarditis group was exclusively male, similar to the other reports of this disease, in which there is a male predominance.^12, 13, 20^ There are significant laboratory differences between classic myocarditis and MIS-C myocarditis, with the MIS-C myocarditis group having higher CRP and higher BNP with more prominent anemia, thrombocytopenia, and lymphopenia, despite a prominent leukocytosis. These findings are congruent with the level of systemic inflammation associated with MIS-C. The etiology of the elevated BNP is unclear, but this finding may be related to the consistent administration of IVIG in the MIS-C cohort, compared to the classic myocarditis group in our study; this volume load may have caused an increase in myocardial stress, reflected by the elevated BNP. In regards to the more significant troponin elevation in the classic myocarditis group, abnormal troponin levels within the first 72 hours have been associated with increased ECMO utilization.^21^ This is similar to what we found in our study in the classic myocarditis group with higher rates of ECMO usage and higher troponin values for this subset, although we used peak troponin values rather than baseline within the first 72 hours.^22^ The patients with vaccine-related myocarditis had similar laboratory studies to the classic myocarditis group, mainly with troponin elevation, without significant laboratory evidence of systemic inflammation.

Notably, all individuals in the COVID-19 vaccine-related myocarditis group that presented with depressed ventricular function regained normal cardiac function at the time of discharge. This finding provides further reassurance that although myocarditis is a possible, albeit rare, sequela of mRNA COVID-19 vaccine, affected patients have recovery of cardiac function in the short term.

### Limitations

There are important limitations in our study. Our data reflect patients hospitalized at a single tertiary pediatric institution. Thus, it represents our center’s approach to classic myocarditis, MIS-C myocarditis, and vaccine-related myocarditis and may not be generalizable to all children with myocarditis. By including only hospitalized patients, we also may have missed patients with subclinical findings, in whom the clinical course may have been different. Additionally, we did not have cardiac MRI data for comparisons between the groups. Given the concerns for active infection and potential hemodynamic instability, patients with MIS-C myocarditis did not undergo cardiac MRI at the time of presentation. Cardiac MRI (cMRI) is the gold standard for non-invasive diagnosis of myocarditis and thus, even in the setting of elevated troponin and symptoms, some patients with MIS-C myocarditis included in this study may not have met cMRI criteria for diagnosis of myocarditis. ^5^ The inclusion of patients with signs of mild myocardial inflammation but not true myocarditis based on cMRI may therefore have overrepresented the number of patients who had full cardiac recovery. The patients with vaccine-related myocarditis did undergo cardiac MRI during their initial hospitalization, but most have not yet undergone follow-up MRI testing. This testing will be important to assess for evidence of ongoing myocardial inflammation and risk for sequelae other than decreased function. Future analyses are planned of the cardiac MRI findings as able in these groups. Finally, three-month outcome data are limited, as some of the patients were lost to follow-up.

## Conclusions

This study highlights the important differences amongst classic myocarditis, MIS-C myocarditis, and COVID-19 vaccine-related myocarditis. Compared to those with classic myocarditis, those with MIS-C myocarditis had more significant hematologic derangements and worse inflammation at presentation, but had better clinical outcomes, including rapid recovery of cardiac function. While patients with COVID-19 vaccine-related myocarditis had similar clinical presentation to patients with classic myocarditis, their pattern of recovery was similar to those with MIS-C, with prompt resolution of symptoms and improvement of cardiac function. These findings are a crucial foundation for healthcare providers, public health officials, and affected patients and families, as knowledge of the cardiac impact of MIS-C and COVID-19 vaccines continues to grow.

## Data Availability

All data referred to in the manuscript is available in our institution's electronic medical record. There is no external data set or supplemental material online that was used.

