## Supplemental Table 1 for "Comparison of MIS-C Related Myocarditis, Classic Viral Myocarditis, and COVID-19 Vaccine related Myocarditis in Children"

Supplemental Table 1: Demographics and Cardiac Symptomology and Signs Among Patients 12 years and older with Classic Myocarditis, MIS-C myocarditis, and Vaccine-related Myocarditis

| **Variable** | **Level** | **N** | **Overall N=88** | **Classic Myocarditis N=32** | **MIS-C N=47** | **Vaccine-related Myocarditis N=9** | **P-Value** |
| --- | --- | --- | --- | --- | --- | --- | --- |
| Age in years, Median (Q1, Q3) |  | 88 | 15.7 (14.1, 16.9) | 16.2 (14.6, 16.9) | 15.1 (13.5, 16.9) | 15.7 (14.5, 16.6) | 0.257 |
| Sex, n (%) | Female | 88 | 17 (19.3) | 7 (21.9) | 10 (21.3) | 0 (0) | 0.300 |
|  | Male |  | 71 (80.7) | 25 (78.1) | 37 (78.7) | 9 (100.0) |  |
| Race/Ethnicity, n (%) | African American | 87 | 43 (49.4) | 16 (50.0) | 26 (56.5) | 1 (11.1) | **0.016** |
|  | Asian |  | 2 (2.3) | 0 (0) | 2 (4.3) | 0 (0) |  |
|  | Hispanic or Latino |  | 14 (16.1) | 5 (15.6) | 9 (19.6) | 0 (0) |  |
|  | Other |  | 6 (6.9) | 3 (9.4) | 1 (2.2) | 2 (22.2) |  |
|  | White |  | 22 (25.3) | 8 (25.0) | 8 (17.4) | 6 (66.7) |  |
| Co-morbidities, n (%) | Asthma | 88 | 11 (12.5) | 5 (15.6) | 4 (8.5) | 2 (22.2) | **0.033** |
|  | Obesity |  | 14 (15.9) | 1 (3.1) | 12 (25.5) | 1 (11.1) |  |
|  | Both |  | 5 (5.7) | 0 (0) | 5 (10.6) | 0 (0) |  |
|  | None |  | 53 (60.2) | 25 (78.1) | 22 (46.8) | 6 (66.7) |  |
|  | Other |  | 5 (5.7) | 1 (3.1) | 4 (8.5) | 0 (0) |  |
| Chest Pain, n (%) | Yes | 88 | 49 (55.7) | 31 (96.9) | 9 (19.1) | 9 (100.0) | **<.001** |
| Dyspnea, n (%) | Yes | 88 | 31 (35.2) | 13 (40.6) | 14 (29.8) | 4 (44.4) | 0.508 |
