## Supplemental Table 2 for "Comparison of MIS-C Related Myocarditis, Classic Viral Myocarditis, and COVID-19 Vaccine related Myocarditis in Children"

Supplemental Table 2: Cardiac Testing Among Patients 12 years and older with Classic Myocarditis, MIS-C myocarditis, and Vaccine-related Myocarditis

| **Variable** | **Level** | **N** | **Overall N=88** | **Classic Myocarditis N=32** | **MIS-C N=47** | **Vaccine-related Myocarditis N=9** | **P-Value** |
| --- | --- | --- | --- | --- | --- | --- | --- |
| Left ventricular ejection fraction at presentation, categorized, n (%) | 01: <30% | 88 | 3 (3.4) | 1 (3.1) | 2 (4.3) | 0 (0) | 0.218 |
|  | 02: 30-55% |  | 45 (51.1) | 15 (46.9) | 28 (59.6) | 2 (22.2) |  |
|  | 03: >55% |  | 40 (45.5) | 16 (50.0) | 17 (36.2) | 7 (77.8) |  |
| Left ventricular ejection fraction at presentation, median (Q1, Q3) |  | 88 | 53.5 (44.1, 61.3) | 55.0 (43.2, 61.0) | 51.6 (44, 61.51) | 59.7 (57.8, 67.2) | 0.085 |
| Worst recorded EF, median (Q1, Q3) |  | 88 | 49 (40, 57.9) | 53.0 (43.5, 60.5) | 45.0 (37.7, 53.0) | 59.7 (57.8, 67.2) | **0.001** |
| Normal EF at the time of discharge, n (%) | Yes | 88 | 76 (86.4) | 24 (75.0) | 43 (91.5) | 9 (100.0) | 0.050 |
| Coronary artery dilation at time of presentation, n (%) | No | 88 | 82 (93.2) | 31 (96.9) | 42 (89.4) | 9 (100.0) | 0.297 |
| Coronary artery dilation at discharge, n (%) | No | 87 | 83 (95.4) | 32 (100.0) | 42 (91.3) | 9 (100.0) | 0.154 |
| Worst pericardial effusion type, n (%) | Absent | 88 | 43 (48.9) | 21 (65.6) | 14 (29.8) | 8 (88.9) | **<.001** |
|  | Present |  | 43 (48.9) | 11 (34.4) | 32 (68.1) | 0 (0) |  |
|  | Trivial |  | 2 (2.3) | 0 (0) | 1 (2.1) | 1 (11.1) |  |
| ECG findings at admission, n (%) | Normal | 87 | 41 (47.2) | 10 (31.3) | 28 (60.9) | 3 (33.3) | 0.070 |
|  | Complete AV block |  | 1 (1.1) | 1 (3.1) | 0 (0) | 0 (0) |  |
|  | Repolarization abnormalities |  | 40 (46.0) | 20 (62.5) | 14 (30.4) | 6 (66.7) |  |
|  | Other^1^ |  | 5 (5.7) | 1 (3.1) | 4 (8.7) | 0 (0) |  |
| ECG findings at clinic follow up (1-2 weeks after discharge), n (%) | Normal  Repolarization abnormalities | 82 | 73 (89.0)  4 (4.9) | 24 (77.4)  3 (9.7) | 44 (97.8)  0 (0) | 5 (83.3)  1 (16.7) | 0.097 |
|  | Other^1^ |  | 5 (6.1) | 4 (12.9) | 1 (2.2) | 0 (0) |  |

Abbreviations: EF (ejection fraction), ECG (electrocardiogram), AV (atrioventricular)

Legend: 1: Other ECG findings included Brugada pattern, atrial tachyarrhythmia, markedly abnormal ventricular axis, junctional tachycardia, and abnormal intervals (i.e. prolonged QTc and PR)
