## Supplemental Table 3 for "Comparison of MIS-C Related Myocarditis, Classic Viral Myocarditis, and COVID-19 Vaccine related Myocarditis in Children"

Supplemental Table 3: Treatment Variation Among Patients 12 years and older with Classic Myocarditis, MIS-C myocarditis, and Vaccine-related Myocarditis

| **Variable** | **Level** | **N** | **Overall N=88** | **Classic Myocarditis N=32** | **MIS-C N=47** | **Vaccine-related Myocarditis N=9** | **P-Value** |
| --- | --- | --- | --- | --- | --- | --- | --- |
| IVIG while admitted, n (%) | Yes | 88 | 58 (65.9) | 18 (56.3) | 39 (83.0) | 1 (11.1) | **<.001** |
| Steroids while admitted, n (%) | Yes | 88 | 54 (61.4) | 9 (28.1) | 45 (95.7) | 0 (0) | **<.001** |
| Vasopressors, n (%) | Yes | 88 | 42 (47.7) | 8 (25.0) | 32 (68.1) | 2 (22.2) | **<.001** |
| Aspirin while admitted, n (%) | Yes | 88 | 63 (71.6) | 20 (62.5) | 42 (89.4) | 1 (11.1) | **<.001** |
| Other NSAID if no aspirin, n (%) | N/A | 85 | 65 (76.5) | 19 (61.3) | 45 (95.7) | 1 (14.3) | **<.001** |
|  | No |  | 3 (3.5) | 1 (3.2) | 2 (4.3) | 0 (0) |  |
|  | Yes |  | 17 (20.0) | 11 (35.5) | 0 (0) | 6 (85.7) |  |
| ECMO, n (%) | Yes | 88 | 5 (5.7) | 3 (9.4) | 2 (4.3) | 0 (0) | 0.464 |
| Mechanical Ventilation, n (%) | Yes | 88 | 7 (8.0) | 4 (12.5) | 3 (6.4) | 0 (0) | 0.399 |
| ICU care during admission, n (%) | Yes | 88 | 48 (54.5) | 8 (25.0) | 38 (80.9) | 2 (22.2) | **<.001** |

Abbreviations: IVIG (intravenous immunoglobulin), NSAID (non-steroidal anti-inflammatory drug), ECMO (extracorporeal membrane oxygenation)
